# Genome-wide association meta-analysis supports genes involved in valve and cardiac development to associate with mitral valve prolapse

**DOI:** 10.1101/2020.07.02.20144774

**Authors:** Mengyao Yu, Sergiy Kyryachenko, Stephanie Debette, Philippe Amouyel, Jean-Jacques Schott, Thierry Le Tourneau, Christian Dina, Russell A Norris, Albert A. Hagège, Xavier Jeunemaitre, Nabila Bouatia-Naji

## Abstract

**Objective:** Mitral valve prolapse (MVP) is a common cardiac valve disease, which affects 1 in 40 in the general population. Previous GWAS have identified six risk loci for MVP. But these loci explained only partially the genetic risk for MVP. We aim to identify additional risk loci for MVP by adding a dataset from the UK Biobank.

**Approaches and Results:** We re-analyzed 1,007/1,469 cases and 479/862 controls from the MVP-France study and the MVP-Nantes study, respectively. We re-imputed genotypes using HRC and TOPMed, and found this latter to perform better in terms of accuracy in the lower ranges of minor allele frequency (MAF) below 0.1. We then incorporated 434 MVP cases and 4,527 controls from the UKBiobank and conducted a meta-analysis GWAS including ∼2000 MVP cases and over 6,800 controls for ∼8 million genotyped or imputed common SNPs (MAF>0.01). We replicated the association on chr2 and now provide a finer association map near *TNS1*. We identified three suggestive risk loci, all driven by common variants on Chr1 (*SYT2*), Chr8 (*MSRA*), and Chr19 (*FBXO46*). Gene-based association using MAGMA revealed 15 risk genes for MVP including *GLIS1, TGFB2, ID2, TBX5, MSRA*, and *DMPK*. Extensive functional annotation showed that genes associated with MVP are highly expressed in cardiovascular tissues, especially heart, and are involved in cardiac development and potentially aging.

**Conclusions:** We report an updated meta-analysis GWAS for MVP using dense imputation coverage and an improved case-control sample. We describe several loci and genes with MVP spanning biological mechanisms highly relevant to MVP, especially during valve and heart development.

TOC category - basic and population studies

TOC subcategory - Arteriosclerosis, Thrombosis, and Vascular Biology

**Highlights:**

- Provide high coverage meta-analysis of GWAS based on TOPMed imputation involving ∼8 million common variants in ∼2000 MVP patients and ∼6800 controls.
- Low frequency variant contributed little to the association of the genes mentioned, where the associations were driven by common variants
- Several association loci involve genes related to cardiac development and potentially aging.

## Introduction

Heart valves are key functional elements of the heart that display specific biological mechanisms in health and disease. During the heart morphogenesis, the mitral valve develops soon after cardiac looping. The complex shape of the mitral valve, with two leaves, allows a very precise balance of force to maintain unidirectional blood flow through the mitral orifice. The importance of valve development in the origin of mitral valve disease has been established through the discovery of the genetic causes of rare syndromes such as Marfan, Loeys-Dietz and Ehlers-Danlos and familial non-syndromic cases.^1,2^ The causal genes play key roles in extracellular matrix deposition and organization, being influenced by TGF beta and/or ciliogenic signaling nodes.

The adult valve can lose its flexibility under the action of permanent mechanical stress and a degenerative process gradually takes place leading to prolapse (MVP), and in many patients, the incapacity of the valve to close. The resulting mitral regurgitation requires surgery repair or replacement, as it greatly increases the risk of heart failure, arrhythmia and even sudden death.^3^ MVP is common, affecting approximately 1 in 40 individuals in the general population.^3^ As for many heart diseases, the establishment of MVP occurs as a result of mild dysfunctions of the many complex biological mechanisms required during development and/or valve function. We have shown through family studies the requirement of *DCHS1*, a member of the cadherin super family, for cell alignment during valve development.^1^ More recently, we have shown that loss of primary cilia during development leads to progressive myxomatous degeneration of the mitral valve in mice and humans.^2^ Using GWAS, we have identified predisposition loci,^1, 4^ particularly those near *TNS1*, are involved in cell adhesion. This finding further supported the importance of cytoskeleton organization revealed by the study of the polyvalvulopathy syndrome caused by *FLNA* mutations.^5^ However, taken together, these loci explain only a small fraction of the genetic contribution to MVP. For instance, the relevance of fine regulation of valve and cardiac development mechanisms is established to be at play for early onset syndromic and non-syndromic valve disease. However, this is unknown for late onset and aging-related valve disease. Identification of additional risk loci will likely provide a better and more complete understanding of the genetic and biological basis of MVP.

One of the limitations of the genetic study of MVP is the lack of large cohorts with genome-wide genotyping that would allow increasing the power to discover new predisposition genes of the current GWAS involving only 1,500 patients.^1^ In addition, since our initial study, it has become possible to achieve much denser and more accurate GWAS through the high-density genetic imputation panels provided by HRC,^6^ and more recently TopMED^7^ consortia. These panels offer a theoretical increase in power, genomic coverage, and fine mapping, notably through the study of low-frequency variants.^7, 8^

In this work, we first compare the imputation performance of the newly generated HRC and TOPMed panels in the context of our cohorts. We thus describe comparable results using these two panels, in favor of a better coverage for low-frequency variants for the TOPMed panel. Then, we performed a GWAS meta-analysis including a new case-control study defined using the UKbiobank resource.^9^ We replicate the *TNS1* locus, and described four new suggestive loci. The gene-based association analysis identifies several new loci that inform established and original mechanisms for the biology underlying the genetic risk for MVP.

## Materials and Methods

### Study populations

We studied 1,920 MVP cases and 6,858 controls in total from three case-control studies from France (MVP-France and MVP-Nantes) and the UK. MVP-France (MVP-F) cases were 1,007 patients recruited through a nation-wide clinical protocol as previously described^1^ (mean age 63.2 years, 68% surgery diagnosis). Approvals were obtained from CPP Ile-de-France VI, (approval n° 60---08, June 25th, 2008), the “Commission Informatique et Libertés” (CNIL) (approval n° 908359, October 14th, 2008) and the French Ministry of Health (ID-RCB: 2008-A00568-47) and was registered on the ClinicalTrial.Gov website (protocol ID: 2008–01). Controls were 1,469 participants ascertained from the Three-City Study (3C), a French multisite population-based study investigating the association between dementia, cognitive impairment and vascular diseases^10^ (mean age 75 years; 79.04% and 10.13% individuals with hypertension and myocardial infarction, respectively). MVP-Nantes (MVP-N) study was described previously^1^ and included 479 French MVP cases (mean age 63 years, 90% surgery diagnosis) and 862 controls (mean age 47 years). The MVP-Nantes study complies with the Declaration of Helsinki, the French guidelines for genetic research and was approved by local ethics committees. Written informed consent was obtained. For the purposes of this study, we ascertained an MVP case-control study from the UK Biobank^9^ (MVP-UKB), which included 434 cases and 4,527 controls. All participants from UK Biobank data were British, Irish and other white backgrounds.^9^ Inclusion criteria for the cases were the main or secondary diagnosis of MVP alone or combined with mitral regurgitation using health records (code = ICD10, I341, I341/I340) and genome-wide genotyping availability. The ethics research committee reference for UK Biobank is 11/NW/0382. We excluded patients with rare syndromes in who MVP usually occurs (e.g., Ehlers-Danlos syndrome and Marfan’s syndrome), and those with ischemic and rheumatic heart diseases. Controls were participants randomly selected among participants without heart disease reported, who were matched to MVP cases for age and sex in a ratio of 1:10.

### Genotyping Data

Genome-wide genotypes were generated in MVP-F study using Illumina Human 660W-Quad and in MVP-N study using Affymetrix Axiom Genome Wide CEU-1. The MVP-UKB study genotypes were obtained using UK Biobank Axiom Array. We applied identical quality control (QC) procedures to the three case-control studies as recommended.^11^ Briefly, individuals with genotyping missing rate < 97%, an excess of ±3 SD of the heterozygosity rate, and with non-European ancestry identified by plotting the first two genotype-driven principal components (PCs) were excluded from further analyses. SMARTPCA was used to identify ancestral outliers^12^ and the background merging data came from HapMap3 (four ethnic populations) and 1000 genomes project (28 ethnic populations). The variants-level QC includes the removal of variants that met SNP missing rate < 97%, MAF < 0.01, and variants deviating from the Hardy-Weinberg disequilibrium (P-value < 10^−6^).

### Haplotype Phasing and Imputation

Imputation was conducted on the Michigan Imputation Server^13^ using minimac3/minimac4.^14^ Both HRC r1.1 2016^6^ and TOPMed Freeze5 on GRCh38^7^ were used as reference panels. Haplotypes phasing was performed using ShapeIT^15^ in the HRC panel imputation and Eagle for the TOPMed panel. We used bcftools (https://samtools.github.io/bcftools/bcftools.html) and vcftools^16^ to check for imputation quality. We excluded variants that did not meet the imputation quality defined as Rsq < 0.3 and MAF < 0.01.

### Statistical analyses

We used SNPTEST^17^ to perform the association analyses (options -frequentist 1 and -method score), which applies logistic regression under the additive model to test the association with MVP status. The first five PCs were used as covariates during the association analyses. Hardy-Weinberg disequilibrium was checked again for variants in cases and controls and we excluded SNPs with P-value < 10^−6^. Meta-analysis was conducted using METAL,^18^ with the option that takes P-values, the direction of effect into account as well as the effective sample size, where Weight = 4/(1/N_cases_+1/N_controls_).

### Genomic risk loci definition and functional annotation

We used FUMA v1.3.4b,^19^ a web-based platform providing diverse post-GWAS annotations to define the genomic risk loci and make the functional mapping of associated variants. As recommended, we defined independent significant SNPs by a GWAS P-value < 1×10^−5^ and moderate correlation (r^2^< 0.6, computed using 1000 genomes Phase 3 data) with any other genome-wide significant SNPs. SNPs in the meta-analysis that were in linkage disequilibrium (LD: r2>0.6) with one of those independent significant SNPs were selected for annotation. In FUMA, the lead SNPs and those in LD with them were annotated using eQTLs from GTEX v8 (selected tissue types: artery aorta, artery coronary, heart atrial appendage, heart left ventricle),^20^ combined annotation dependent depletion (CADD v1.4)^21^ and RegulomeDB (RDB v1.1).^22^ RNA *in situ* hybridization in mouse embryo for Tgfb2, Id2, Tbx5, Smg6, Srr, Abcc3, Six5, Dmwd and Syt2 at E14.5 was obtained through Genepaint (https://gp3.mpg.de, accessed in June 2020).^23, 24^

Regional association plots were created using LocusZoom.^25^ Positions change between genome assembly GRCh38/hg38 and GRCh37/hg19 was completed using Liftover and Kaviar.^26^

### MAGMA gene-based association and gene-set analysis

The SNPs and P-values from the meta-analysis were used as the inputs for the gene-based association and gene-set analysis using MAGMA v1.07 in FUMA.^19, 27^ MAGMA combines the P-values of SNPs that are within window sizes of 50kb upstream and 50kb downstream of the genes and takes LD between SNPs into account. We used 19,151 protein-coding genes from the NCBI 37.3 gene definitions, giving a Bonferroni-corrected P-value threshold of 2.611 × 10^−6^.

We also used MAGMA to analyze 9,996 predefined gene sets (GO terms obtained from MsigDB v7.0).^28^ The competitive analysis was used to test whether the combined effects of genes in a gene set are strongly associated with MVP than the combined effects of all other genes not in the gene set.

## Results

### Imputation accuracy between HRC and TopMED in MVP GWAS datasets

We first aimed to assess the usefulness of using larger imputation reference panels in the two French MVP case controls studies. MVP-France study (MVP-F) included 1,007 cases and 1,469 controls with 492,438 genotyped variants. MVP-Nantes study (MVP-N) included 479 cases and 862 controls with 370,697 genotyped variants. We generated 229,973,672 and 230,053,813 imputed variants in MVP-F and MVP-N, respectively, from the TOPMed reference panel, which represent 5.8-fold more variants compared to HRC (39,117,105 in MVP-F and MVP-N) (Figure 1a). However, most of these variants had an imputation score Rsq smaller than 0.3: 84% (MVP-F-TOPMed), 88% (MVP-N-TOPMed), 51% (MVP-F-HRC), and 60% (MVP-N-HRC) (Figure 1a). Compared to MVP-N, the relatively larger sample size of MVP-F study generates a slightly larger number of well-imputed variants (Figure 1a).

**Figure 1.**
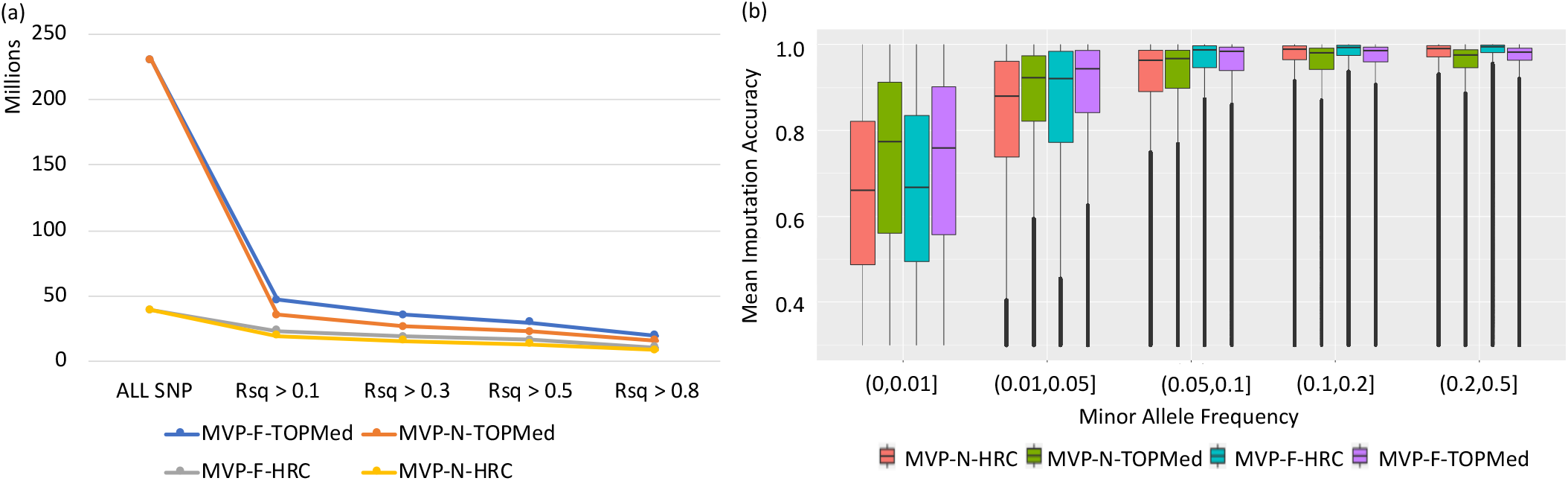
Comparison of the imputation quality of the results for MVP-F and MVP-N using HRC and TOPMed as reference panel, respectively. (a). The numbers of variants generated using different imputation accuracy thresholds; (b). The imputation quality at different MAF region using all the cleaned SNPs (Rsq > 0.3) in cohorts MVP-Paris and MVP-Nantes. Rsq: imputation accuracy; MVP-F: MVP-France cases-control study. MVP-N: MVP-Nantes case-control study.

We then classified MAF into five regions ((0,0.01], (0.01,0.05], (0.05,0.1], (0.1,0.2] and (0.2,0.5]). Most of the variants with Rsq < 0.3 are rare variants which minor allele frequency (MAF) is smaller than 0.01, as illustrated for all SNPs generated on chromosome 22 (Supplementary figure I). We also found that TOPMed panel imputed variants present, on average, a better accuracy for low frequency ranges (0,0.01], (0.01,0.05] and (0.05,0.1], while relatively stable accuracy at MAF ranges (0.1,0.2] and (0.2,0.5] compared to HRC panel in both studies (Figure 1b). Taken together, we use TOPMed as the imputation panel to allow analyzing more well imputed variants, especially in the low frequency (0.01<MAF<0.1) category.

### Genetic association analyses in French and UKBiobank case control studies

We meta-analyzed three GWAS involving a total of 1,920 MVP cases and 6,858 controls and ∼8 million (8,021,974) genotyped or imputed common SNPs (MAF>0.01). We confirmed a deviation from the expected levels of significance in this updated meta-analysis (Figure 2a).

**Figure 2.**
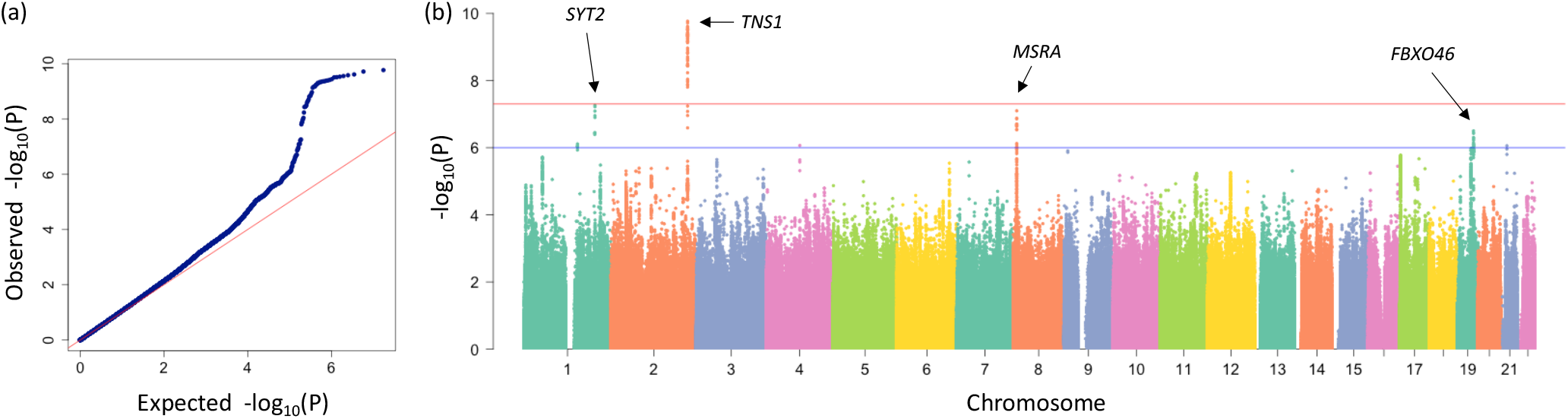
SNP-based association results with MVP in a GWAS meta-analysis involving MVP-F, MVP-N and MVP-UKB case-control studies (lambda GC = 1.08). (a) The Q-Q plot represents the expected (x-axis) versus the observed (y-axis) P-values; (b) Manhattan plot summarizes the -log10 (P) of each SNP by chromosome obtained from the GWAS meta-analysis. The blue line indicates the threshold for suggestive association (P-value < 1 × 10^−6^) and the red line indicates the genome-wide significance threshold (P < 5 × 10^−8^). Both plots represent the association of ∼ 8 million SNPs genotyped and imputed using TOPMed panel.

The strongest association signal was observed on chromosome 2 at the *TNS1* locus, where we report 46 variants with a genome-wide significant P-value < 5×10^−8^ (Figure 2b). In addition to successfully replicating the previously reported top associated SNP rs12465515 (P-value = 2.8×10^− 10^; OR = 1.30[1.19-1.42]), we found a new top associated variant on chromosome 2 (lead SNP: rs7595393; P-value = 1.71×10^−10^; OR = 1.31[1.20-1.42], Table 1, Supplementary figure II). Both SNPs are highly correlated (r^2^=95, 1000G Phase 3: CEU). As expected, conditional analyses both on rs12465515 and rs7595393 at the *TNS1* locus resulted in disappearance of the association signal (Figure 3). Functional annotation of 103 SNPs in LD (r2>0.6) with 4 independent significant SNPs (ISS) at this locus indicated the existence of 5 SNPs predicted to be deleterious alleles (CADD score > 12) and 7 SNPs likely to lie within regulatory elements according to the RDB score (RDB score > 3a) (Supplementary Table I).

**Table 1.**
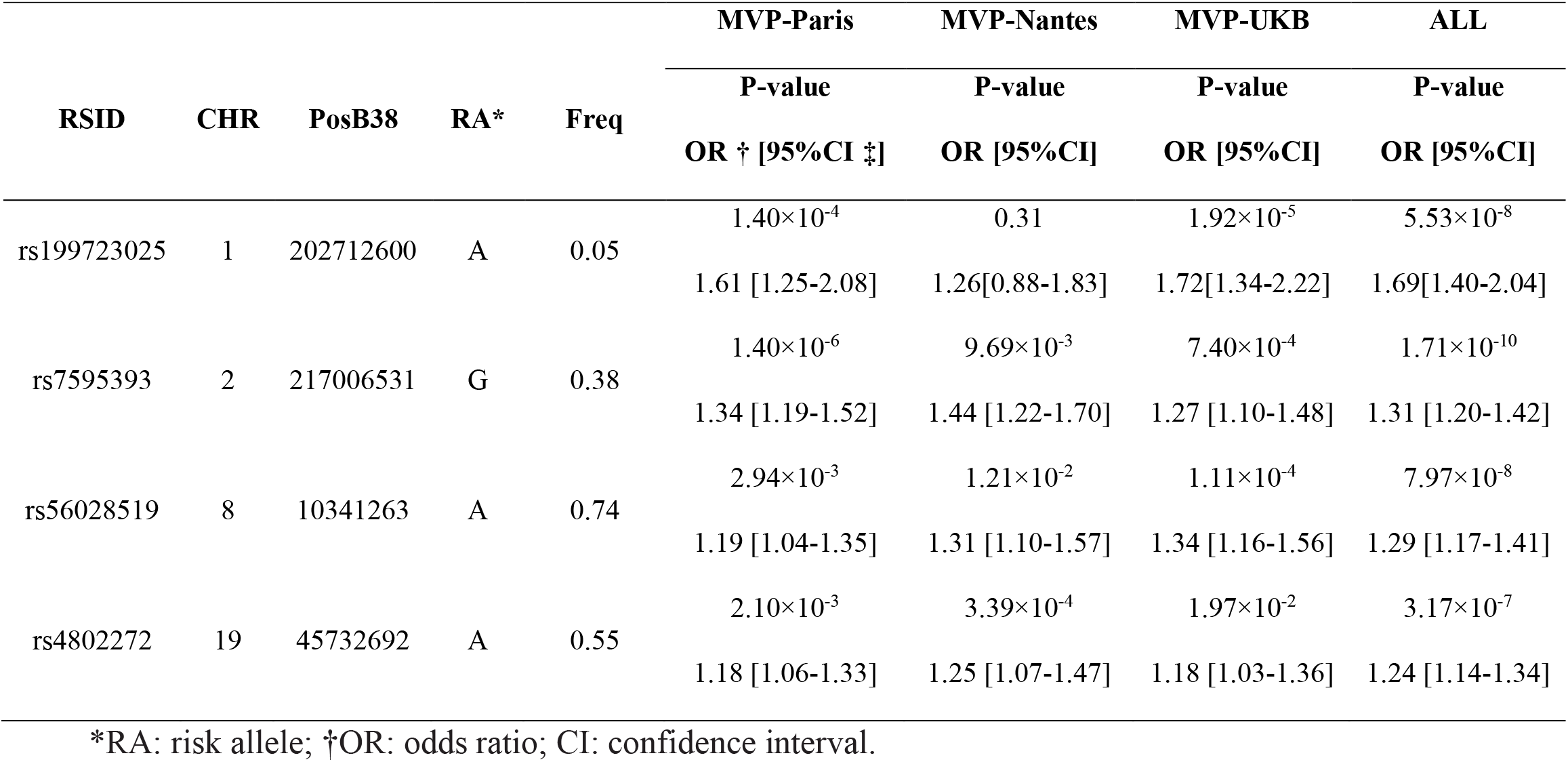
Associations of top SNPs with MVP obtained in the GWAS meta-analysis.

| RSID | CHR | PosB38 | RA* | Freq | MVP-Paris | MVP-Nantes | MVP-UKB | ALL |
| --- | --- | --- | --- | --- | --- | --- | --- | --- |
|  |  |  |  |  | P-value | P-value | P-value | P-value |
|  |  |  |  |  | OR † [95%CI ‡] | OR [95%CI] | OR [95%CI] | OR [95%CI] |
| rs199723025 | 1 | 202712600 | A | 0.05 | 1.40×10<sup>-4</sup> | 0.31 | 1.92×10<sup>-5</sup> | 5.53×10<sup>-8</sup> |
|  |  |  |  |  | 1.61 [1.25-2.08] | 1.26[0.88-1.83] | 1.72[1.34-2.22] | 1.69[1.40-2.04] |
| rs7595393 | 2 | 217006531 | G | 0.38 | 1.40×10<sup>-6</sup> | 9.69×10<sup>-3</sup> | 7.40×10<sup>-4</sup> | 1.71×10<sup>-10</sup> |
|  |  |  |  |  | 1.34 [1.19-1.52] | 1.44 [1.22-1.70] | 1.27 [1.10-1.48] | 1.31 [1.20-1.42] |
| rs56028519 | 8 | 10341263 | A | 0.74 | 2.94×10<sup>-3</sup> | 1.21×10<sup>-2</sup> | 1.11×10<sup>-4</sup> | 7.97×10<sup>-8</sup> |
|  |  |  |  |  | 1.19 [1.04-1.35] | 1.31 [1.10-1.57] | 1.34 [1.16-1.56] | 1.29 [1.17-1.41] |
| rs4802272 | 19 | 45732692 | A | 0.55 | 2.10×10<sup>-3</sup> | 3.39×10<sup>-4</sup> | 1.97×10<sup>-2</sup> | 3.17×10<sup>-7</sup> |
|  |  |  |  |  | 1.18 [1.06-1.33] | 1.25 [1.07-1.47] | 1.18 [1.03-1.36] | 1.24 [1.14-1.34] |

\*RA: risk allele; †OR: odds ratio; CI: confidence interval.

**Figure 3.**
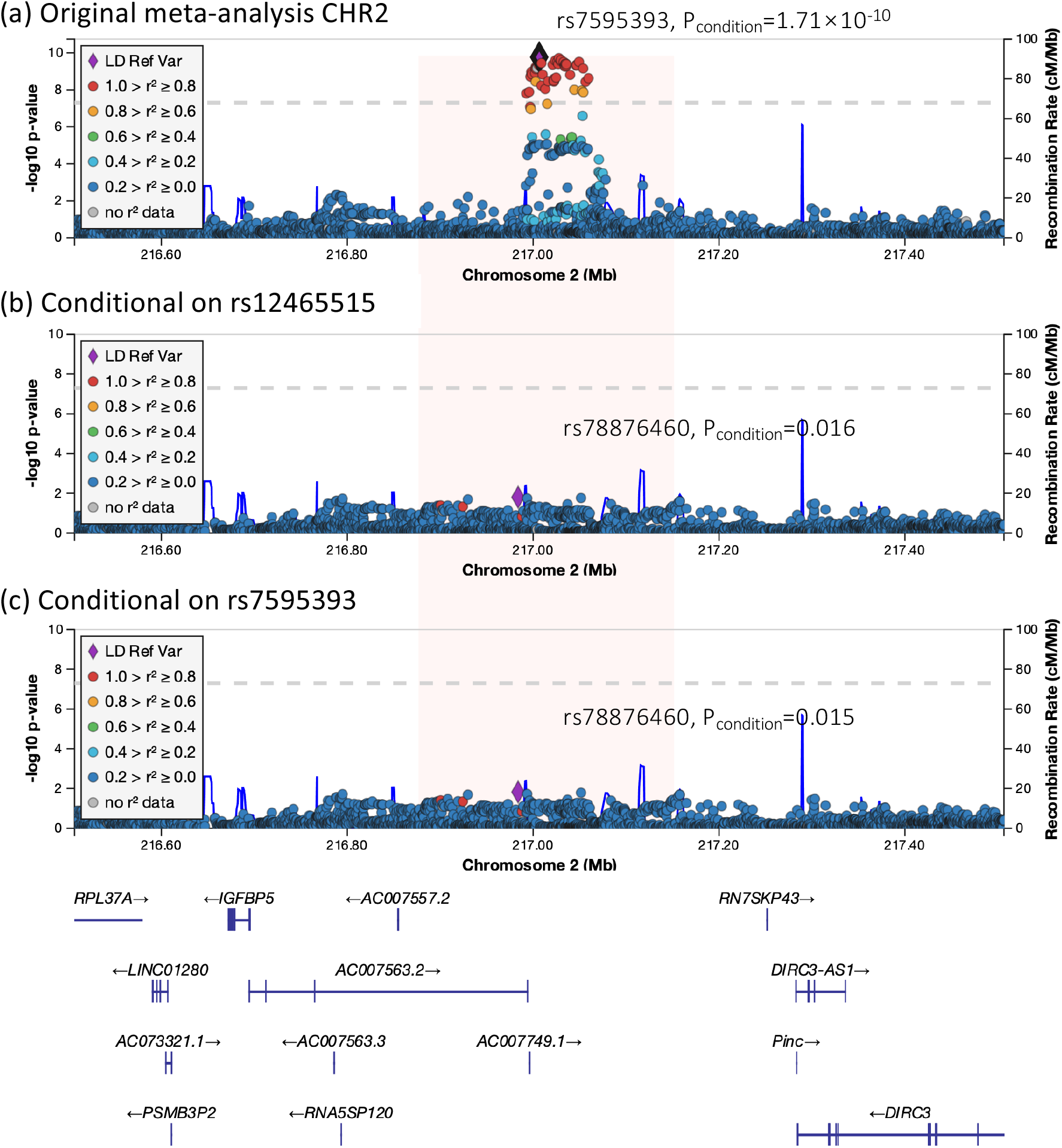
Conditional Analysis of the *TNS1* locus. Locus Zoom plots of the *TNS1* locus demonstrate conditional analysis of Dina reported lead SNP rs12465515, the top SNP rs7595393 using GCTA-COJO. Linkage disequilibrium of SNPs with the conditioned SNPs is based on data from MVP-F cohort and is shown by the color of the points. The best signals in each plot are marked as a purple rhombus. (a) Association of SNP with MVP; (b) MVP association conditioned on Dina reported lead SNP rs12465515 (rs12465515 added as an additional covariate); (c) MVP association conditioned on the top SNP rs7595393

We also report three suggestive and original association signals (Figure 2, Table 1). On chromosome 1, the lead SNP (rs199723025, effect allele frequent (EAF) = 0.05, P-value = 5.53×10^−8^, OR = 1.69[1.40-2.04]) is an intergenic deletion variant of Synaptotagmin 2 gene (*SYT2*) (Table 1, Supplementary figure III). We found that the lead variant is a significant eQTL in atrial appendage and artery aorta (P-value: 2.4×10^−5^ and 1×10^−8^) for the lysine demethylase 5B gene (*KDM5B*) involved in DNA stability and repair. The second locus on chromosome 8 (rs56028519, EAF = 0.74, P-value = 7.97×10^−8^, OR = 1.29[1.17-1.41]) is an intronic variant of the methionine sulfoxide reductase A gene (*MSRA*) (Table 1, Supplementary figure IV). The lead SNP rs56028519 is an eQTL of a LncRNA gene (AF131215.2) in heart atrial appendage (P-value=4.5×10^−7^) and a LncRNA gene (RP11-981G7.6) in left ventricle (P-value=1.3×10^−7^). We found that several SNPs in LD (r^2^>0.6) with the 3 ISS are located near the regulatory elements, especially rs11249991 and rs11781529 that showed high potential to be regulatory (RDB score > 3a). Three SNPs in total are likely to be deleterious, including rs11783281 (CADD score = 19.05, Supplementary Table I). The third suggestive association signal was located in a particularly gene-rich region on chromosome 19 (rs4802272, EAF = 0.55, P-value = 3.17×10^−7^, OR = 1.24[1.14-1.34]) (Table 1, Supplementary figure V). Although the lead SNPs mapped to *FBXO46*, the association signal spans five genes in total, including, *SIX5, FOXA3, RSPH6A, DMPK* and *DMWD* (Supplementary figure 5). The lead SNP is an eQTLs of *DMPK* and *DMWD* in artery aorta (P-value: 2.1×10^−9^, 3.4×10^−11^). Two SNPs in high LD with the lead SNP showed high deleteriousness scores (rs62111759, CADD score = 12.93 and rs672348, CADD score = 12.43) (Supplementary Table I).

### Genome-wide gene-based association and pathway analyses

We performed a genome-wide gene-based association analysis using MAGMA^27^ to estimate the gene level association on the basis of all SNPs in a gene. This method is a complementary approach to single SNP GWAS analyses and usually reveals genes with locally consistent associated SNPs that individually may do not reach genome-wide significance. In total, 15 genes reached gene-wide significant association (MAGMA P-value < 2.611×10^−6^) with MVP (Figure 4a, 4b). We highlight *GLIS1* (P=2.50×10^−6^) that we have previously identified to associate with MVP using single SNP and pathway analyses,^4^ *TGFB2* (P=1.74×10^−6^), a high-profile candidate gene for myxomatous valve disease, *MSRA* (P= 5.18×10^−7^), one of the suggestive loci in the SNP GWAS, and *TBX5* (P = 1.10×10^−6^), a key regulator of heart development (Table 2). The ten remaining genes mapped into two gene-rich loci on chromosome 17 and 19. On chromosome 17, five genes reached gene-wide significance including *SMG6* (P-value = 1.06×10^−7^), *SRR* (P-value = 6.17×10^− 9^), in addition to *TSR1* (P-value = 1.04×10^−8^), *SGSM2* (P-value = 3.98×10^−8^) and *ABCC3* (P-value = 1.10×10^−6^) (Table 2). On chromosome 19, we report *FBXO46* (P-value = 2.05×10^−6^), *SIX5* (P-value = 9.25×10^−9^), *DMPK* (P-value = 6.18×10^−8^), *DMWD* (P-value = 9.86×10^−8^) and *RSPH6A* (P-value = 6.42×10^−7^) (Table 2). Tissue expression analyses using the GTEx data resource showed that four of genes (*DMPK, DMWD, TBX5* and *ID2*) are strongly expressed in cardiovascular tissues, especially in the heart atrial appendage and heart left ventricle (Figure 4c). We note that low frequency variant contributed little to the association of the genes above mentioned, where the associations were driven by common variants (Supplementary Figure VI). Section *in situ* hybridization validated developmental expression of many of these gene within the mitral valves (Supplementary Figure VII) further supporting their potential involvement in disease phenotype.

**Table 2.** Genes significantly associated with MVP in gene-based association tests (GWGAS) using MAGMA.

| CHR | START | STOP | NSNPS* | ZSTAT | P-value | SYMBOL |
| --- | --- | --- | --- | --- | --- | --- |
| 1 | 53921910 | 54249877 | 827 | 4.5652 | $2.50 \times 10^{-6}$ | *GLIS1* |
| 1 | 218469577 | 218667961 | 420 | 4.6398 | $1.74 \times 10^{-6}$ | *TGFB2* |
| 2 | 8768975 | 8874583 | 332 | 4.8734 | $5.49 \times 10^{-7}$ | *ID2* |
| 8 | 9861778 | 10336401 | 2023 | 4.8846 | $5.18 \times 10^{-7}$ | *MSRA* |
| 12 | 114741736 | 114896247 | 527 | 4.7335 | $1.10 \times 10^{-6}$ | *TBX5* |
| 17 | 1913133 | 2257065 | 895 | 5.1892 | $1.06 \times 10^{-7}$ | *SMG6* |
| 17 | 2156677 | 2278554 | 358 | 5.6949 | $6.17 \times 10^{-9}$ | *SRR* |
| 17 | 2175797 | 2290801 | 330 | 5.6058 | $1.04 \times 10^{-8}$ | *TSR1* |
| 17 | 2190792 | 2334352 | 391 | 5.368 | $3.98 \times 10^{-8}$ | *SGSM2* |
| 17 | 48662138 | 48819613 | 358 | 4.7339 | $1.10 \times 10^{-6}$ | *ABCC3* |
| 19 | 46163887 | 46284162 | 227 | 4.606 | $2.05 \times 10^{-6}$ | *FBXO46* |
| 19 | 46218043 | 46322484 | 182 | 5.6254 | $9.25 \times 10^{-9}$ | *SIX5* |
| 19 | 46222975 | 46335810 | 235 | 5.2882 | $6.18 \times 10^{-8}$ | *DMPK* |
| 19 | 46236205 | 46346060 | 265 | 5.2019 | $9.86 \times 10^{-8}$ | *DMWD* |
| 19 | 46248968 | 46368577 | 332 | 4.8423 | $6.42 \times 10^{-7}$ | *RSPH6A* |

Note: GWGAS tests were performed in MAGMA and represent the aggregate effect of all SNPs in a gene (defined by GRCh37 reference genome locations). The Bonferroni threshold of significance for 19,151 genes was $2.611 \times 10^{-6}$ . The position in the table is B37. \*NSNPS: the number of SNPs annotated to that gene that were found in the data and were not excluded based on internal SNP QC.

**Figure 4.**
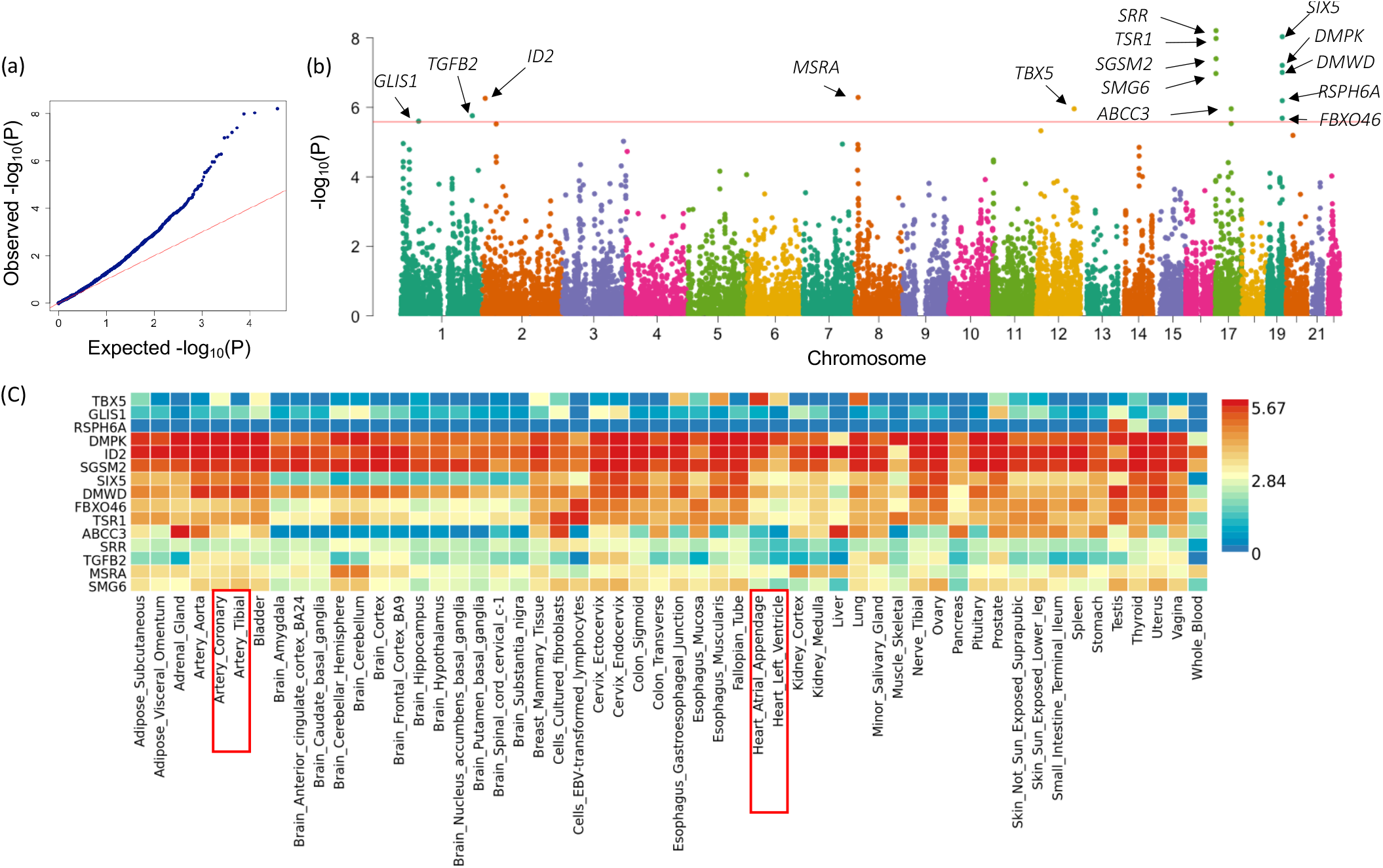
Genome-wide gene-based association analysis and the heatmap of expression of the highlighted genes. Genome-wide gene-based analysis of 19,151 genes that were tested for association with MVP using MAGMA. (a) Q-Q plot showing the expected (x-axis) versus the observed (y-axis) P-values. (b) Manhattan plot summarizing the -log10(P) of each gene ordered by chromosome. The red line indicates the Bonferroni corrected threshold for genome-wide significance (P = 0.05/19151= 2.611×10^−5^). The top 15 most significant genes are labeled (Clone-based (Vega) gene and RNA Genes are not labeled). (c) The heatmap of tissue expression of the 18 highlighted genes using the GTEx v8 54 tissue types data resource.

MAGMA pathway analysis using 9,996 gene sets (GO terms obtained from MsigDB) indicated that several significantly enriched gene sets for MVP associated genes are related to cardiac biology. We highlight cardiac ventricle formation (P-value = 3.09×10^−6^), cardiac chamber formation (P-value = 8.94×10^−6^), cardiac right ventricle morphogenesis (P-value = 1.11×10^−4^), cardioblast differentiation (P-value = 1.24×10^−4^), apoptotic process involved in heart morphogenesis (P-value = 1.81×10^−4^), atrial septum morphogenesis (P-value = 3.08×10^−4^), cardiac atrium development (P-value = 3.12×10^−4^) and a GO term related to ciliary rootlet (P-value = 4.05×10^−4^) (Supplementary table II).

## Discussion

Here we describe a high genetic coverage meta-analysis of GWAS based on TOPMed imputation involving ∼8 million common variants in ∼2000 MVP patients and ∼6800 controls. In addition to replicate the association at the *TNS1* locus, we identified several associated variants and genes involving in established and original mechanisms for the biology underlying the genetic risk for MVP.

Our results using the TOPMed imputation panel provided higher resolution association map for the risk of MVP in a reasonably well powered dataset. We provide confirmatory results and fine mapping at the *TNS1* where we now report rs7595393 as the new lead associated SNP. The association signal on Chr2 is located in a gene-desert genomic region. We have previously provided solid biological evidence, which included a myxomatous valve phenotype in the heterozygote knockout mouse supporting the gene encoding Tensin 1, a focal adhesion protein, to be causal.^1^ Functional annotations at this locus describe several SNPs belonging to the association block of rs7595393 as plausible causal variants. In the absence of eQTL, functional annotation for open chromatin and enhancer marks specifically in the mitral valve, we acknowledge that *in silico* annotation has limited ability to point at the causal variants and further experiments are needed to achieve this goal. One important addition in the current study is the improved coverage of low frequency variants (0.01<MAF<0.10), Our current data do not allow us to firmly conclude about the putative role of low frequency variants in MVP genetic risk, which needs to be explored further using larger datasets. However, the fact that the single-SNP and gene-based association signals we report were overwhelmingly driven by common variations does not support this hypothesis.

In addition to confirming the association on Chr2, we describe three unprecedented suggestive association signals in *SYT2* (Chr1), *MSRA* (Chr8) and *FBXO46* (Chr19) that will need to be replicated in larger studies. Two of these association signals (*MSRA* and *FBXO46* loci) were further supported by the gene-based association results. We also report gene association for *GLIS1* and several genes at the *SMG6/SRR* locus where we have previously described single SNP associations with MVP.^1, 4^

Several newly associated genes with MVP are involved in established biological mechanisms and are highly relevant to mitral valve disease. This applied to the TGF-beta family member *TGFB2*, an essential growth factor for myocardial cells endothelial to mesenchymal transition and valve elongation during valve development.^29^ It has been established that the myxomatous degeneration is the result of the response of endothelial cells to mechanical stress, aging, or excessive stimulation of TGF beta proteins.^29^ Both in the Marfan mouse model^30^ and human mitral valves,^31^ TGF beta induces pro-fibrotic function and promotes the proliferation and differentiation of endothelial cells into myofibroblasts.

Our gene-based association results strengthen the pivotal role that may play regulatory genes involved in cardiac development in susceptibility to MVP. This is supported by the association of inhibitor of DNA binding 2 gene (*ID2*) involved in cell differentiation and proliferation, which action is regulated by TGF-beta1 and BMP-7.^32^ Interestingly, *Id2* expression is lost in valve forming regions of Smad4-deficient endocardium in mice.^33^ Moreover, Id2 was shown to belong to the same molecular pathway that includes Tbx5, that coordinates ventricular conduction system lineage in mice.^34^ *TBX5* association with genetic risk for MVP is intriguing. *TBX5* is a member of the T-box transcription factor and key regulator of cardiac development, specifically involved in cardiac conduction and maintenance of mature cardiomyocyte.^35^ Mutations in *TBX5* cause the Holt-Oram syndrome (OMIM #142900), a disorder characterized by abnormalities in upper limbs and congenital heart lesion. *TBX5* is also an established risk locus identified from GWAS for traits related to cardiac conduction, mainly atrial fibrillation,^36^ QRS duration^37^ and PR interval.^38^ *TBX5* expression is not detected in human heart valves,^39^ which may suggest the valve prolapse phenotype to result from defaults in the interplay between myocardium, conduction tissue and mitral valve apparatus during heart development or later as a consequence of valve aging.

Our genetic study also describes original mechanisms that may deserve future biological investigation. *MSRA* encodes methionine sulfoxide reductase A, a ubiquitously expressed and conserved enzyme, including in the heart, with the highest levels in the kidney and the nervous system. Several GWAS association signals near *MSRA* were reported for blood pressure,^40^ neuroticism,^41^ and glomerular filtration.^42^ The biological implication of MSRA in the degenerative process of the valve is unclear, and could be through this enzyme protective role against oxidative stress during aging.^43^

Our study presents several limitations. The inclusion of a new dataset from the UKBiobank and the generation of a dense association map did not compensate the limited power of our study and the original suggestive loci described will need to be replicated in future studies. The gene-based results are not able to detect association signals involving gene-desert genomic regions with long-range enhancers, as the one we observe on Chr2 upstream *TNS1*. Another limitation of this method is that gene-rich genomic regions provides redundant association signals involving the same sets of variants and do not allow to point specifically at a potential causal gene. Functional annotation is based on existing eQTLs and gene expression pattern in cardiovascular tissues, especially heart atrial appendage and left ventricle, where cell composition and gene expression may differ from gene expression in the mitral valve.

## Conclusions

To summarize, we report an updated meta-analysis GWAS for MVP using dense imputation coverage and an increased case control sample. We describe several established and original associated loci and genes with MVP spanning biological mechanisms highly relevant to heart valve disease. Follow-up biological studies in cell and animal models are needed to better understand their direct effect on the valve degenerative process.

## Data Availability

Data is available to nonprofit organization researchers upon request from the corresponding author.

## Acknowledgments

We acknowledge the contribution of the Leducq Foundation, Paris for supporting the genetic study in the French case control studies. We thank Carolina Roselli and Patrick Ellinor for providing access to TopMED panel data. This research has been conducted using the UK Biobank Resource under Application Number 32360.

## Abbreviations

MVP: Mitral Valve Prolapse
HRC: Haplotype Reference Consortium
TOPMed: Trans-Omics for Precision Medicine
SNP: Single Nucleotide Polymorphism
GWAS: Genome-Wide Association Study
eQTLs: Expression Quantitative Trait Locus
MAF: Minor allele frequency
RA: Risk Allele
OR: Odds Ratio
CI: Confidence Interval

